# Single-cell transcriptomics predicts relapse in *MLL*-rearranged acute lymphoblastic leukemia in infants

**DOI:** 10.1101/2020.04.14.20056580

**Authors:** Tito Candelli, Pauline Schneider, Patricia Garrido Castro, Luke A. Jones, Rob Pieters, Thanasis Margaritis, Ronald W. Stam, Frank C.P. Holstege

## Abstract

Infants with *MLL*-rearranged acute lymphoblastic leukemia (ALL) undergo intense therapy to counter a highly aggressive leukemia with survival rates of only 30-40%. The majority of patients initially show therapy response, but in two-thirds of cases the leukemia returns, typically during treatment. Accurate relapse prediction would enable treatment strategies that take relapse risk into account, with potential benefits for all patients. Through analysis of diagnostic bone marrow biopsies, we show that single-cell RNA sequencing can predict future relapse occurrence. By analysing gene modules derived from an independent study of the gene expression response to the key drug prednisone, individual leukemic cells are predicted to be either resistant or sensitive to treatment. Quantification of the proportion of cells classified by single-cell transcriptomics as resistant or sensitive, accurately predicts the occurrence of future relapse in individual patients. Strikingly, the single-cell based classification is even consistent with the order of relapse timing. These results lay the foundation for risk-based treatment of *MLL*-rearranged infant ALL, through single-cell classification. This work also sheds light on the subpopulation of cells from which leukemic relapse arises. Leukemic cells associated with high relapse risk are characterized by a smaller size and a quiescent gene expression program. These cells have significantly fewer transcripts, thereby also demonstrating why single-cell analyses may outperform bulk mRNA studies for risk stratification. This study indicates that single-cell RNA sequencing will be a valuable tool for risk stratification of *MLL*-rearranged infant ALL, and shows how clinically relevant information can be derived from single-cell genomics.

**Key Points:**

- Single-cell RNA sequencing accurately predicts relapse in *MLL*-rearranged infant ALL
- Identification of cells from which *MLL*-rearranged infant ALL relapses occur

## Introduction

Acute lymphoblastic leukemia (ALL) in infants (i.e. children <1 year of age) is frequently driven by chromosomal translocations of the *Mixed Lineage Leukemia* (*MLL* or *KMT2A*) gene, which occur in ∼80% of the cases. Translocations of the *MLL* gene on chromosome 11q23 lead to fusions of the N-terminus of *MLL* to the C-terminus of one of many known translocation partner genes. Although >90 translocation partner genes have been identified, the majority of the infant ALL patients carry one of three recurrent types of *MLL* translocations in which the *MLL* gene becomes fused to either *AF4* (aka *AFF1*; 49% of the cases), *ENL* (aka *MLLT1*; 22% of the cases), or *AF9* (aka *MLLT3*; 16% of the cases)^1^. *MLL*-rearranged infant ALL represents a rare but highly aggressive type of childhood leukemia that is notoriously characterized by chemotherapy resistance and high relapse rates, leading to a very poor prognosis. Regardless of the type of *MLL* translocation, event-free survival (EFS) rates for *MLL*-rearranged infant ALL patients remains at 30-40%, when treated according to the international collaborative INTERFANT treatment protocol^2,3^. In contrast, with EFS chances of approximately 75%, infant ALL patients not carrying an *MLL* translocation fare significantly better^1-3^, thereby approaching the superior clinical outcome of childhood ALL patients older than 1 year of age, for whom current EFS rates currently reach 90%^4,5^.

Despite the massive disparity in EFS, the majority (∼95%) of *MLL*-rearranged infant ALL patients initially do achieve disease remission. Within the first weeks of induction therapy, most patients show rapid reductions in leukemia burden. In two-thirds of the cases, however, the leukemia reemerges, typically within the first year from diagnosis and while still on treatment, giving rise to an even more chemotherapy-resistant form. Not surprisingly, once a relapse has occurred, the survival chances are even further reduced and the majority of such patients succumb to the leukemia^6^. High mortality is not the only problem. Patients who survive this malignancy, face significant additional challenges. Due to the aggressive nature of this malignancy, standard treatment for *MLL*-rearranged infant ALL patients is very intense in these young patients^2,3,7^. The prolonged exposure to various toxic substances during infant development may lead to detrimental side effects, including long-term adverse effects in survivors at a later stage in life^8^. Further intensification of the current chemotherapeutic treatment is not an option, due to increased risks of therapy-related toxic mortality. There clearly is ample room for improving treatment of *MLL*-rearranged infant ALL.

A crucial first step towards increasing survival and reducing side-effects simultaneously, would be accurate risk assessment, preferably at diagnosis. Accurately predicting which patients are at high-risk of leukemia relapse, and subsequent identification and characterization of the nature of the relapse-initiating cells, would allow the development of therapies that prevent disease reemergence, and at the same time may allow the reduction of therapy intensity for patients that are unlikely to experience disease relapse. Many hypotheses about relapse emergence involve cellular heterogeneity^9-15^, and a high degree of clonal heterogeneity has been observed in *MLL*-rearranged infant ALL^16,17^. Given the transformative ability of single-cell genomics to analyze heterogeneous systems^18-23^, we decided to study *MLL*-rearranged infant ALL relapse occurrence by single-cell RNA sequencing (scRNA-seq).

## Methods

### Patient samples

Bone marrow (BM) biopsies taken at diagnosis were from infants (<1 year) with ALL and treated according to the international collaborative INTERFANT treatment protocols^2,3^. Samples were either from patients with at least seven-year relapse-free survival or from patients who experienced early relapse (within 1 year after diagnosis). All samples used were from *MLL*-rearranged pro-B infant ALL patients, carrying either of the two most common *MLL* fusion genes, i.e. *MLL*-*AF4* or *MLL*-*ENL*^24^. Care was taken to spread attributes such as sex and translocation type across the dataset (Table 1). Informed consent was obtained from the parents or legal guardians according to the Helsinki declaration. BM biopsies were processed and frozen viably as described^25^. Leukemic blast percentages (Table 1) were determined microscopically using May-Grünwald Giemsa stained cytospin preparations.

**Table 1:** Patient characteristics.

| Sample ID | **1977N** | **1702N** | **635N** | **8010R** | **6806R** | **4662R** | **4483R** |
| --- | --- | --- | --- | --- | --- | --- | --- |
| Translocation | t(4;11) | t(11;19) | t(4;11) | t(4;11) | t(4;11) | t(11;19) | t(11;19) |
| Gender | female | male | male | female | male | female | male |
| Age at diagnosis(months)<sup>1</sup> | 6.5 | 11.1 | 2.8 | 0 | 11.3 | 5.3 | 3.6 |
| Time to relapse(months)<sup>2</sup> |  |  |  | 7.0 | 10.3 | 4.8 | 4.6 |
| Risk stratification<sup>3</sup> | medium | medium | high | medium | medium | high | high |
| Lymphoblasts (%)<sup>4</sup> | 92 | 95 | 92 | 98 | 99 | 98 | 94 |

<sup>1</sup>Days after birth divided by 30.
<sup>2</sup>No value means no relapse for the duration of follow-up (minimally 7 years).
<sup>3</sup>Risk stratification according to Interfant<sup>1,31</sup>. For *MLL-r* iALL there is only medium and high risk.
<sup>4</sup>Lymphoblasts in bone marrow sample used for single-cell sequencing.

### Fluorescence-activated cell sorting (FACS)

Samples were thawed and resuspended in 1xPBS, 2mM EDTA, 0.5%BSA and 2.5 μg/mL 7AAD or 5 μg/mL DAPI (BD Biosciences) in a concentration of approximately 1 million cells/mL. Viable single cells were sorted based on forward/side scatter properties and 7AAD/DAPI staining using FACS (FACSAria III, BD Biosciences) into 384-well plates (Biorad) containing 10 µL mineral oil (Sigma) and 50 nL of barcoded reverse transcription primers as described^26^ (Supplemental Table 1). Data was acquired using FACSDiva version 8.0.1, the gating strategy employed for sorting is shown in supplemental Figure 1.

### FACS metrics

Index sorting files containing cytometric data for each sorted cell were recovered for all sequenced plates. This allowed association between transcriptomic data and forward scatter area values as depicted in Figure 3. To obtain forward scatter area values for larger populations of cells, Flow Cytometry Standard (FCS) files for each of the samples were analysed. These samples were gated by the same strategy used for sorting into plates (Supplemental Figure 1), yielding cytometric data on a minimum of 4224 cells (sample 635N) and maximum of 170866 cells (sample 8010R). When comparing forward scatter area values across groups of patients (Figure 3E, aggregate) an identical number of cells (n=4224) were taken from each patient to ensure equal representation.

### Single-cell RNA sequencing (scRNA-seq)

384-well plates with sorted cells were processed into Illumina sequencing libraries as described^26,27^. Libraries were paired-end sequenced at 75 bp read length using the Illumina NextSeq500 sequencer. Reads were aligned to human genome version GRCh38.p10, GENCODE release 26, with ERCC external RNA control sequences appended using STAR version 2.6.1^28^. Read assignment was performed using the “comprehensive gene annotation for main chromosomes” annotation file obtained from GENCODE GRCh38.p10 release 26. Reads were processed into count-tables as described^29^. Because of their high variation in gene expression, at this stage mitochondrial genes were removed. Failed reactions were identified by low levels of ERCC external RNA controls and excluded^29^. In order to distinguish live cells from dead and/or apoptotic cells, a liveness threshold was calculated for each plate based on the wells with no cell added^29^. This threshold was set to a minimum of 500 transcripts. After filtering, analysis was performed using R version 3.3.4 and the package Seurat^30^ version 2.1.0 with default parameters unless stated otherwise. Per-cell transcript counts were normalized to 3500 transcripts. The first 15 principal components (PCs) of a PC analysis calculated using the expression of highly variable genes were used to generate t-distributed stochastic neighbour embedding (t-SNE) plots (Figure 1b-c, Supplemental Figure 2) and perform Louvain clustering^30^ (Figure 1c) using a resolution of 1.1. Cluster number 9 consisted of T-cells (Supplemental Figure 2a-b) and was excluded from further analyses.

**Figure 1:**
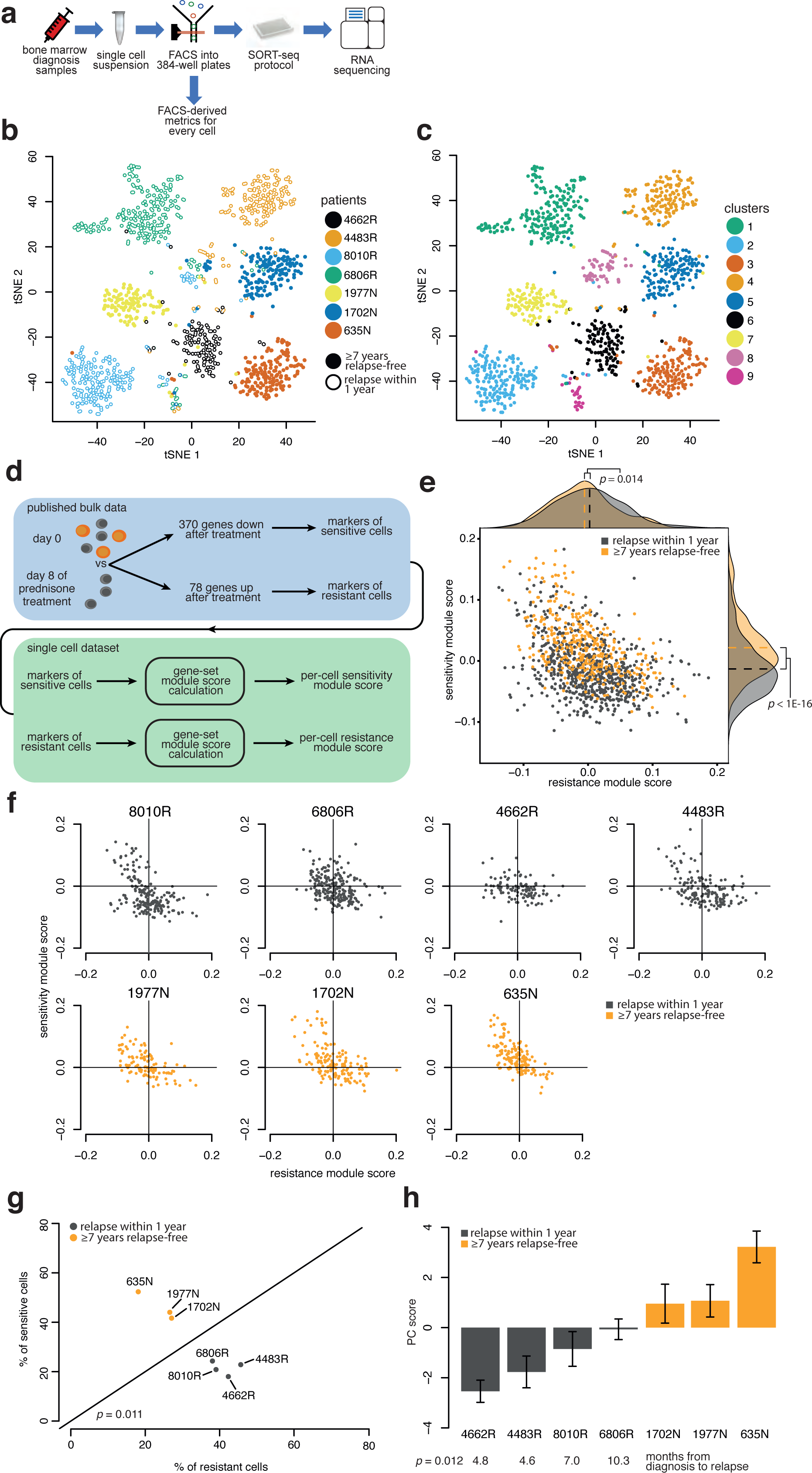
Single-cell drug-sensitivity classification leads to relapse prediction. **a**, Experiment design. **b**, t-distributed stochastic neighbour embedding (t-SNE) plot of cells labelled according to sample ID, with R indicating patients who suffered relapse and N no relapse. c, Louvain clustering^30^ projected onto the t-SNE plot. Clusters 8 and 9 correspond to T-cells and actively dividing cells respectively (Extended SupDataplementary Figure 1a-d). **d**, Previously published bulk mRNA data of an apparent prednisone gene expression response in patients^32^ was applied as gene modules to classify cells for sensitivity (down-regulated genes) and resistance (up-regulated genes). This is based on the interpretation that some cells are resistant (grey) and some sensitive to prednisone (orange). **e**, Gene module scores (x- and y-axis) for each cell, with cells from patients who later developed relapse labelled grey and cells from relapse-free patients orange. **f**, Gene module scores for cells from each patient individually. Cells in the upper-left quadrant are predicted to be more sensitive and in the bottom-right more resistant to treatment. **g**, Quantification of the fraction of cells from each patient (from f) predicted to be sensitive (y-axis) or resistant (x-axis). **h**, First principal component (PC) score from analysis of genes from the two modules for cells from each patient. Error-bars represent standard error of the mean.

### Cell-cycle analysis

Cell-cycle phase (Figure 3c, Supplemental Figure 2) was determined for each cell using the Seurat^30^ (version 2.1.0) CellCycleScoring function with default parameters. The list of marker genes for S and G2M phase are as described^31^.

### Gene module scores

For sensitivity and resistance module scores (Figure 1-2, Supplemental Figure 3-5), the differentially expressed genes were obtained, as described^32^. Genes upregulated at day 8 (78 genes) were considered markers for therapy-resistance, while genes downregulated after treatment (370 genes) were considered markers of sensitivity (Figure 1d). Calculation of module scores was performed using the Seurat^30^ (version 2.1.0) AddModuleScore function with the following modification: when sampling control genes, the same gene can be selected multiple times. This ensures that the distribution of values is centred around zero and eliminates biases due to uneven number of genes when two modules are compared.

### Principal Component (PC) score

The PC score constitutes the first PC of a PC analysis calculated using the union of sensitivity and resistance module genes (see gene module score above). As depicted in Supplemental Figure 3, a high PC score corresponds to cells with a predicted high sensitivity to treatment.

### *In vitro* prednisolone treatment

*In vitro* drug exposures were performed^33^ by incubation with 100 µg/mL prednisolone (BUFA), the liver-activated form of prednisone, or with vehicle for 3 days. Cells were viably frozen^25^ and later thawed for scRNA-seq. The blast percentage was determined as described above to ensure that each sample had at least 90% blasts before being processed.

### Differential expression analyses

To find markers of the cell clusters depicted in Figure 1c, differentially expressed genes (used in Supplemental Figure 2a) were determined with the FindAllMarkers function in Seurat^30^. Differential expression was assessed using the bimodal test (argument test.use = “bimod”, only.pos = TRUE). The resulting *p*-values were Bonferroni multiple-testing corrected. Genes with an adjusted *p*-value lower than 0.05 and with an average log fold-change (natural log) above 0.25 were considered differentially expressed. This resulted in 271 cluster-8 specific genes and 330 cluster-9 specific genes. To determine genes differentially expressed between sensitive and resistant cells (Figure 3a), two groups were composed consisting of the 15 cells with the lowest PC score from each patient (resistant cells) and the 15 cells with the highest PC score from each patient (sensitive cells). These two groups were compared using the function FindMarkers with the same arguments as above. To eliminate the effect of patient-specific gene expression, we excluded all genes to which a single patient contributed more than 40% of the total number of cells expressing that gene. Genes exceeding the thresholds described above were considered differentially expressed (Supplemental Table 2).

### Gene Ontology (GO) enrichment

GO category enrichment was calculated using the compareCluster function from the clusterProfiler R package^34^, see *supplemental methods* for details.

### Cell size determination by microscopy

Images of May-Grünwald Giemsa stained cytospin slides were made using the DM200 LED microscope (Leica) and utilized to create outlines of the cells to determine cell size using ImageJ software^35^. See *supplemental methods* for a more detailed description.

## Results

### Clustering of leukemic cells according to individual patients

To identify subpopulations of cells potentially associated with relapse, approximately 3000 leukemic cells were analysed with over 350 million scRNA-seq reads. Samples were standard bone marrow biopsies taken at initial diagnosis, derived from patients who remained relapse-free for at least seven years, or from patients who relapsed within 1 year from diagnosis. The samples cover the two most recurrent *MLL* translocations found among infant ALL patients, including t(4;11) and t(11;19), giving rise to the MLL fusion genes *MLL-AF4* and *MLL-ENL*, respectively^1-3^. Patient characteristics are listed in Table 1, including the current risk assessment^2,3^ which is known to be insufficiently accurate. Samples were processed with a modified version of CEL-Seq2^27^ (Figure 1a), a medium throughput scRNA-seq platform that provides high sensitivity^36^ and cytometric data on individual cells. As anticipated, clustering of cells is largely according to individual patients (Figure 1b-c). This agrees well with the personalized nature of cancers^37^ and the substantial patient to patient heterogeneity of *MLL*-rearranged infant ALL^16,17^. Taking the leukemic blast percentages of >90% for all samples used in this study (Table 1) into account, implies that the cells clustering according to individual patients are the leukemic cells. Two additional small clusters occur with contributions from multiple patients (Figure 1b-c, clusters 8-9). These two smaller clusters correspond to T-cells, and a subpopulation of more actively dividing cells respectively (Supplemental Figure 2a-d). When analysed in this unsupervised manner, the clusters of leukemic cells do not group by characteristics such as translocation type or relapse occurrence (Supplemental Figure 2f-h). This clearly underscores the distinct nature of individual cancers and the challenge of predicting treatment outcome, and could explain why current risk assessment is far from accurate.

### Single-cell drug-sensitivity classification leads to prediction of relapse and relapse-timing

To deal with gene expression variation in bulk mRNA measurements, supervised approaches have previously been applied to training and validation cohorts to find clinically relevant gene expression signatures^38^. Here a classification framework was devised that is specific for scRNA-seq data. The approach employs an independently derived external dataset and is devoid of supervised training, focusing initially on a key functional characteristic rather than on clinical outcome immediately. In the majority of *MLL*-rearranged infant ALL cases, relapses arise during treatment^2,3^. ALL therapy includes the cornerstone glucocorticoid drug prednisone. The response to this drug has been studied by a variety of approaches, including bulk mRNA measurements in samples derived from pediatric ALL patients^32^. Rather than interpreting such data as revealing a prednisone gene expression response, we reasoned that genes showing apparent upregulation may actually represent a signature of cells already resistant to treatment beforehand. Similarly, genes with apparent downregulation may represent markers of cells sensitive to prednisone and therefore disappear during treatment. Based on this rationale (Figure 1d), we directly applied the prednisone gene expression response^32^ as modules for therapy resistance (78 upregulated genes) and sensitivity (370 downregulated genes). This enabled classification of individual cells as being more sensitive or more resistant to therapy, based on the expression of the two modules (Figure 1e). The distribution of cells is a continuum from apparent sensitivity to apparent resistance. Strikingly, upon labelling of cells according to whether or not a future relapse occurs, a significant difference is observed, in particular for the sensitivity module (*p*<10^−16^) that is more strongly expressed in cells from patients who remain relapse-free. Treatment resistance is an obvious determinant of outcome^39^. These analyses indicate that this property can be predicted for individual cells using scRNA-seq and that such a single-cell classification may predict relapse potential.

To further test the predictive capability of our data, the single-cell classification was next examined in individual patients (Figure 1f). Visual inspection indicates more resistant-predicted cells (Figure 1f, bottom-right quadrants) in patients who eventually relapse and more sensitive-predicted cells (upper-left quadrants) in patients who remain relapse-free. For quantitative comparison, the percentage of cells classified as sensitive/resistant is calculated for each diagnostic sample. This yields a strong distinction between patients with and without relapse (Figure 1g). Strikingly, even the order of relapse-timing is correctly predicted, as is evident from the ordering of patients by the first principal component derived from the two gene modules combined (Figure 1h, see Supplemental Figure 3a-b for how well the first principal component embodies the signal from the two modules).

### *In vitro* prednisolone treatment enriches for cells classified as resistant

The single-cell classification is based on the idea that the gene expression response to prednisone^32^ reflects survival of treatment-resistant cells (Figure 1d). To further test this, untreated leukemic cells from diagnosis were exposed to prednisolone, the liver-activated form of prednisone, *in vitro* (Figure 2a). Single-cell classification shows that leukemic cells predicted to be resistant are present in a lower proportion in the control sample and become highly enriched after elimination of the sensitive cells by prednisolone (Figure 2b-e). This agrees with the interpretation that the previously published prednisone response genes are markers for treatment sensitivity/resistance (Figure 1d).

**Figure 2:**
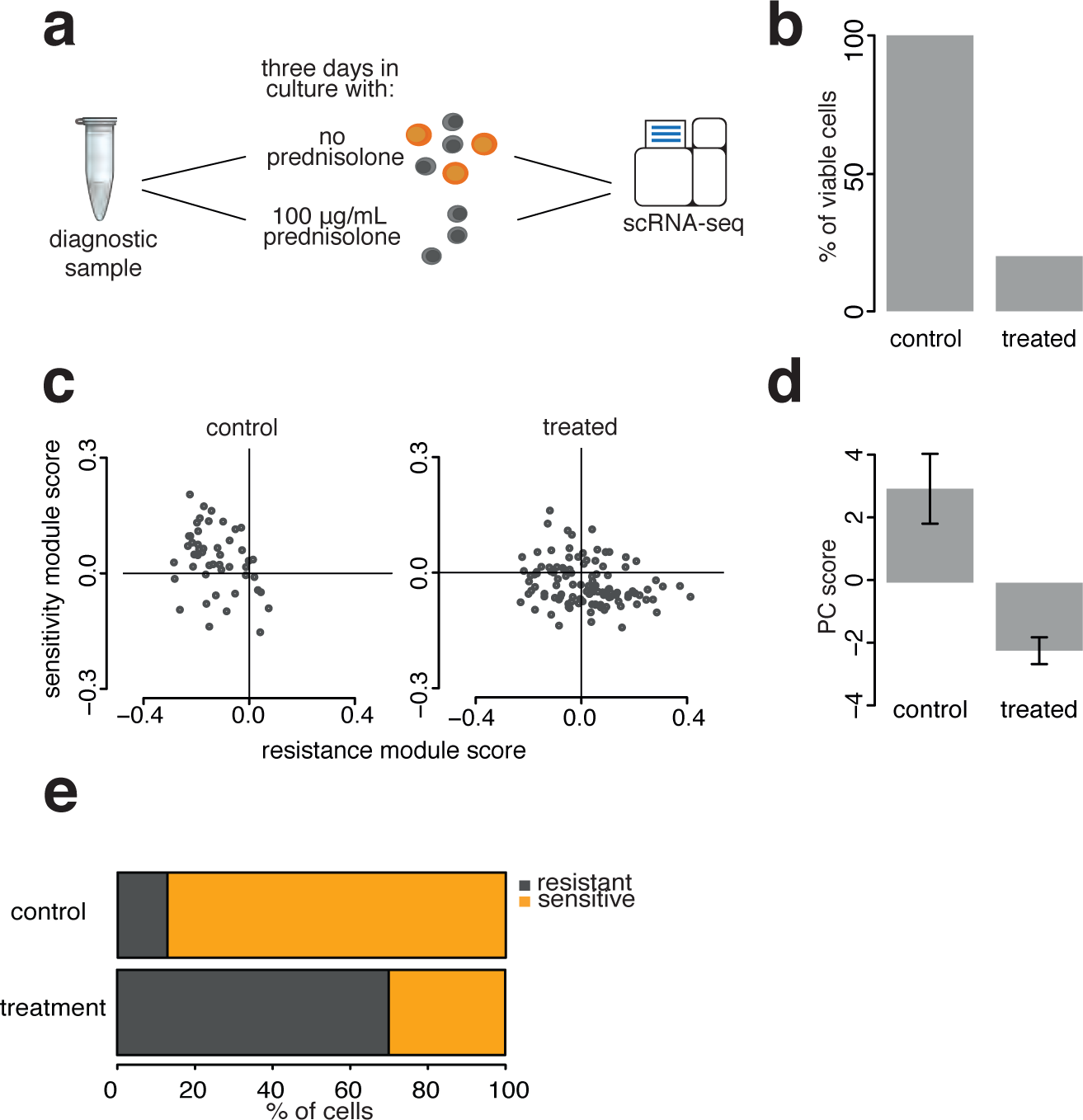
*In vitro* treatment enriches for cells classified as resistant. **a**, Untreated leukemic cells from bone marrow diagnostic biopsy were cultured with and without prednisolone, the liver-activated form of prednisone. **b**, Cell viability after treatment. **c**, scRNA-seq based sensitivity and resistance module scores of viable cells from control and treated cultures as in Figure 1f. d. First PC score as in Figure 1h. e. Fractions of cells classified as sensitive/resistant in control and treated samples.

### Cells associated with high relapse risk share properties with quiescent/dormant cells

High proportions of cells classified as therapy-resistant at diagnosis, are associated with higher relapse risk (Figure 1-2). It is obviously important to understand the nature of such cells. Although leukemic cells from individual patients are distinct (Figure 1b), the classification shows that they can also manifest as a continuum when analysing the sensitivity/resistance gene modules (Figure 1e). To further characterize sensitive/resistant cells, exhaustive differential expression analysis was performed. The expression pattern of significantly up-regulated genes in predicted resistant/sensitive cells also indicates a continuum of characteristics rather than distinct subtypes (Figure 3a). Gene Ontology indicates that cells with predicted higher sensitivity to treatment are more actively dividing (Figure 3b). This is supported by Seurat^30^ gene expression cell-cycle analysis (Figure 3c), that also emphasizes the converse: therapy resistant-predicted cells are associated with reduced cell-cycle activity.

**Figure 3:**
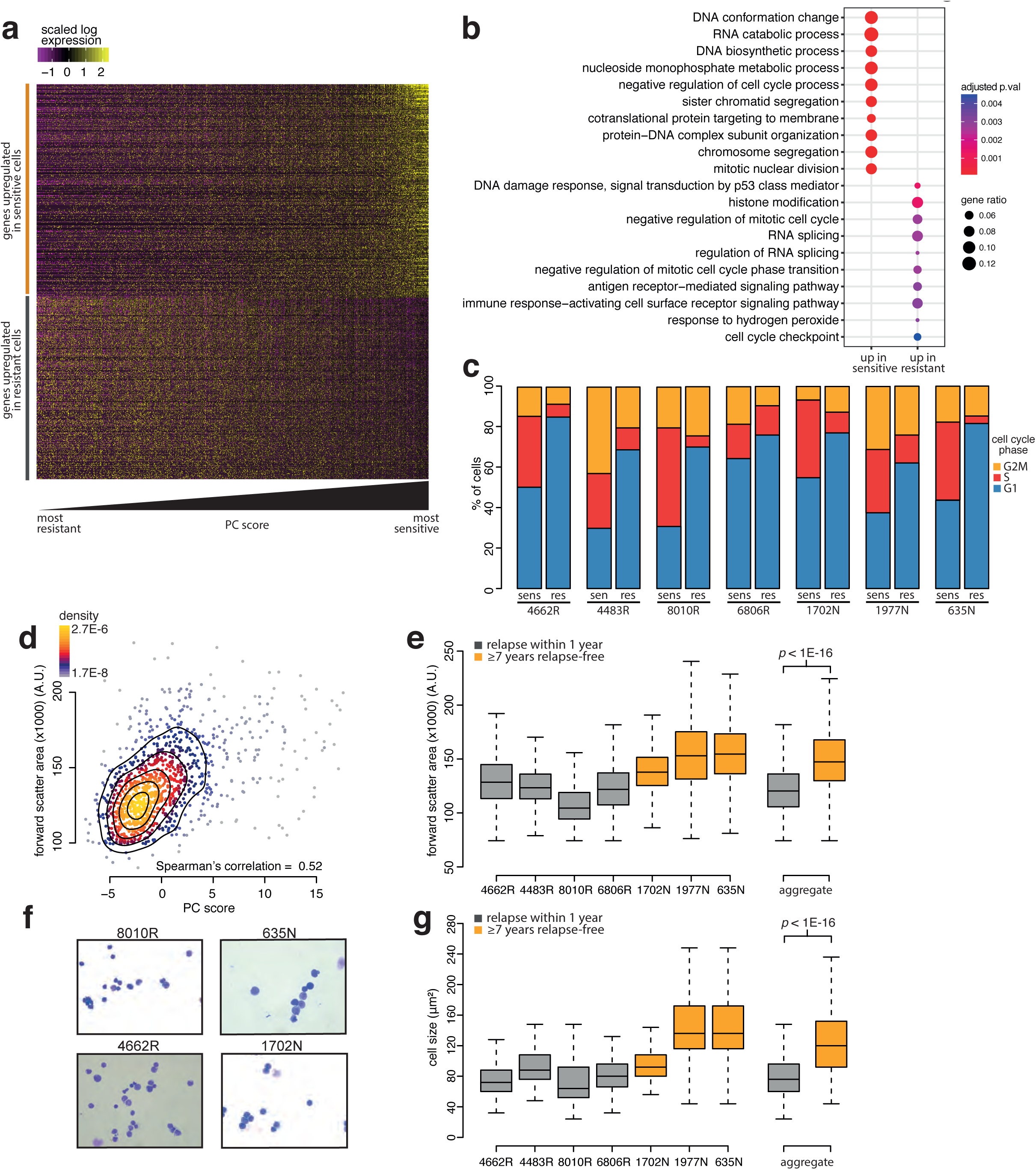
Cells associated with high relapse risk share properties with quiescent/dormant cells. **a**, Expression heatmap of all cells, sorted left to right on the first PC score. The genes depicted are those upregulated in cells predicted to be either sensitive (331 genes) or resistant (282 genes), listed in Supplemental Table 1. **b**, Gene Ontology categories enriched in the upregulated genes in sensitive and resistant cells. Gene ratio represents the fraction of differentially expressed genes in each category **c**, Seurat^30^ cell cycle analysis of cells predicted to be sensitive or resistant (Figure 1f) in the different samples. **d**, Correlation between FACS forward scatter area and the first PC score of individual cells analysed by scRNA-seq. **e**, Distribution of forward scatter area values of all cells from individual samples by box plots. Each samples contribution to the aggregates was equal. **f**, Examples of cytospin microscopy images used to determine the cell sizes depicted in **g**, Cell size determination by quantification of images (Extended DataSupplemental Figure 3b).

### Cells from relapsing patients have a smaller average size at diagnosis

Cells with quiescent/dormancy properties have previously been implicated in resistance to cancer treatment^9-15^. Besides low cell-cycle activity, another hallmark of quiescent cells is smaller size. To independently ascertain whether cells associated with increased relapse risk may indeed be quiescent/dormant, cell size was investigated. The scRNA-seq technology used here applies FACS to sort individual cells prior to processing (Figure 1a). FACS forward light scatter is often applied as a proxy for cell size and there is indeed an association between the forward scatter area of the cells analysed by scRNA-seq and the first principal component of the sensitivity/resistant gene modules (Figure 3d-e). This is also confirmed directly by quantification of cell size by imaging (Figure 3f-g, Supplemental Figure 4). Compared to cells from patients who remain free from relapse, cells from patients who do relapse have a smaller average size before treatment starts (Figure 3d-g).

### Lower amounts of transcripts in relapse-associated cells hampers classification by bulk mRNA expression analyses

Beyond indicating the cells from which relapse arises and the potential for improving treatment of these vulnerable patients, this study also reveals why single-cell analyses may in some cases outperform bulk mRNA approaches for patient classification. The smaller cells associated with higher risk of disease relapse, have substantially lower numbers of transcripts (Supplemental Figure 5a). This fits with quiescence/dormancy and means that bulk mRNA data will not proportionately represent the relative abundance of such cells. Indeed, applying the gene modules (Figure 1) on previously published bulk mRNA *MLL*-rearranged infant ALL datasets does not result in a relapse/non-relapse distinction (Supplemental Figure 5b). Bulkifying the scRNA-seq data by complete pooling of all transcripts yields a dataset that also does not discriminate well (Supplemental Figure 5c). However, pooling the scRNA-seq data after down-sampling so that each cell contributes an equal number of transcripts, does yield “bulk” data on which the modules discriminate between patients who do and do not relapse (Supplemental Figure 5d).

## Discussion

To date, *MLL*-rearranged infant ALL remains an aggressive and difficult to treat childhood malignancy. Although induction therapy leads to complete remissions in the vast majority of cases (∼95%), two-thirds of the patients experience disease relapse within one year from diagnosis, while treatment is still ongoing^2,3^. This suggests that most of the leukemic cells are responsive to treatment, while small subpopulations of therapy-refractory cells exist from which the leukemia re-emerges, despite ongoing therapy. Hence, clinical outcome largely depends on the absence or presence of such therapy-resistant leukemic cells, and future improvements in treatment should involve the identification of chemotherapeutics able to eliminate these cells.

In order to confirm the existence of therapy-refractory and relapse-initiating cells at diagnosis, we here utilized single-cell RNA sequencing to investigate relapse occurrence in *MLL*-rearranged infant ALL. Our data underlines the high degree of heterogeneity of *MLL*-rearranged infant ALL, also observed by others^16,17^. Unsupervised analysis shows that individual leukemic cells cluster to individual patients, rather than to other characteristics such as clinical outcome, type of *MLL* translocation, or age at diagnosis. Despite the need to classify patients into subtypes and stratify risk assessment based on patient characteristics, this clearly shows that every patient is also unique.

Current risk stratification of *MLL*-rearranged infant ALL involves categorising patients into either being medium-risk or high-risk patients, based on age at diagnosis, white blood cell counts, and the *in vivo* response to 7 days of prednisone treatment. Patients currently classified as high-risk are <6 months of age, have white blood cell counts at diagnosis of 300 × 10^9^/L or more, and show a poor prednisone response. All other *MLL*-rearranged infant ALL patients are categorised as medium-risk. Although this division does lead to significant differences in clinical outcome^2,3^, it is still often inaccurate, as shown in Table 1. Based on classic risk stratification criteria, one of the samples in this study was determined to be high-risk but remained event-free for at least 7 years, whereas two of the patients in this study who experienced an early relapse (within 1 year from diagnosis) were determined to be at medium-risk. A possible explanation for this may lie in some of the criteria by which patients currently are being categorized as medium or high risk, i.e. high white blood cell counts and an initially poor response to prednisone. The latter parameter dictates that the response to prednisone of a given patient is poor when 7 days of prednisone treatment fails to drop the leukemic blast percentage beneath a defined threshold. Although this measurement certainly could be indicative for the proportion of cells that is highly sensitive to prednisone or to chemotherapy in general, it does not necessarily provide a measure for the small amounts of truly therapy-resistant cells that evade treatment all together. With the majority (∼95% of the cases) of *MLL*-rearranged infant ALL patients reaching complete remission upon induction therapy, without detectable minimum residual disease (MRD), these subsets of therapy-resistant cells presumably are very small. Therefore, a parameter determining the amount of these therapy-resistant cells at diagnosis may well outperform current risk stratification.

Based on the scRNA-seq data, we show that relapse occurrence can already be predicted at initial diagnosis by quantifying the scRNA-seq classification of individual cells as either being therapy resistant or sensitive. Importantly, this is achieved using a completely independent dataset^32^, with no training on outcome, but rather by classifying individual cells for an autonomous property that is a likely determinant of clinical outcome. Furthermore, this approach not only shows the ability to accurately predict whether or not patients would experience a relapse, but was also able to predict the timing of the relapse, albeit on a very small group of patients. Interestingly, this stratification model solely relies on the identification of the percentage of treatment-resistant leukemic cells present at diagnosis, regardless of the type of *MLL* translocation, white blood cell counts, or age at diagnosis. Moreover, these analyses indicate a continuum of properties, rather than a distinct subtype of cells. Patients who present with a higher proportion of cells at the resistance end of the continuum evidently have a higher relapse risk.

The independent dataset^32^ used to categorize *MLL*-rearranged infant ALL cells into treatment sensitive or resistant, was based on differences in gene expression in samples from older children with ALL, before and after 7 days of prednisone mono-therapy. Yet, we reasoned that the cells identified by scRNA-seq to be at the resistance end of the continuum, truly represent cells resistant to therapy in general, rather than cells only resistant to prednisone. In concordance with this assumption, earlier published gene expression signatures specifically associated with the *in vitro* response to prednisolone (the liver-activated metabolite of prednisone) in pediatric ALL^40^ or more specifically in *MLL*-rearranged infant ALL^41^ do not overlap with the gene expression profile identified in the present study.

The identified therapy-resistant cells associated with higher risk of disease relapse typically represent small and non-proliferating cells that, based on the associated gene expression profile, clearly share properties with quiescent/dormant cells. These smaller quiescent leukemic cells have substantially lower numbers of transcripts and are therefore not proportionately represented in bulk data. This all agrees with the idea that signals from cell types with reduced transcript levels are challenging to identify robustly within patient-derived bulk mRNA data. Single-cell analyses resolve this and may therefore enable subclassification of other disease-states that, like *MLL*-rearranged infant ALL, are influenced by the abundance of subpopulations of cells with different transcript numbers. As demonstrated, the approach for patient classification based on predicting an independent, functional attribute of individual cells by scRNA-seq, and applying this quantitatively, provides a powerful framework for enabling such studies.

Taken together, these results are consistent with models whereby relapse arises from a subpopulation of pre-existing, therapy-resistant cells. Furthermore, these data demonstrate how single-cell based risk assessment may, in future, fulfil the unmet need of more accurate risk stratification. On one hand, this may lead to the reduction of the intensity of the treatment, and therefore reduce adverse side-effects for patients that are unlikely to experience disease relapse. At the same time this will oblige us to invest into further characterization of the identified treatment-resistant, quiescent/dormant leukemic cells and develop immuno- and targeted therapies very specifically directed against these cells in particular. Eventually, elimination of these therapy-resistant cells during early phases of the treatment may well prevent relapse occurrence in a substantial number of cases, leading to increased survival.

## Data Availability

Datasets generated for this study have been deposited in EGA, accession number pending.

## Data availability

Datasets generated for this study have been deposited in EGA under accession number xxx (submission has started and the acc. nr. will be included in a final version).

## Code availability

Any custom code used in this study is available upon request.

## Ethical approval

All samples were collected at the Erasmus MC – Sophia Children’s Hospital. Ethical approval was provided by the Erasmus MC Institutional Review Board.

## Acknowledgements

This work is supported by KiKa and the European Research Council (ERC) advanced grant 671174. We thank the Princess Máxima Center and the Hubrecht Institute core FACS facilities for support, as well as members of the Holstege, Kemmeren and Stam groups at the Princess Máxima Center for advice and discussions.

## Author contributions

T.M., R.W.S. and F.C.P.H. conceived and designed the study; R.W.S. and F.C.P.H. arranged funding; P.S., P.G.C., T.M., R.W.S. and F.C.P.H. selected samples; P.S., P.G.C., R.W.S. and R.P. obtained samples; P.S., P.G.C. and T.M. processed samples and performed experiments; P.S., P.G.C. and L.J. provided data; T.C., P.S., P.G.C., T.M. and R.W.S. processed and analysed data; R.P. helped with clinical interpretation; all authors contributed to interpreting results; T.C. and P.S. made figures; T.C., P.S., T.M., R.W.S. and F.C.P.H. wrote the first draft; all authors verified and/or corrected the final version and contributed to writing; T.C. deposited data; R.W.S. and F.C.P.H. supervised the project.

